# The impact of heating, ventilation, and air conditioning design features on the transmission of viruses, including the 2019 novel coronavirus: a systematic review of ultraviolet radiation

**DOI:** 10.1101/2021.10.12.21264904

**Authors:** Gail M. Thornton, Brian A. Fleck, Natalie Fleck, Emily Kroeker, Dhyey Dandnayak, Lexuan Zhong, Lisa Hartling

**Author notes:** **Correspondence:** Dr. Brian A. Fleck; Department of Mechanical Engineering, Faculty of Engineering, University of Alberta; 116 Street and 85 Avenue; Edmonton, Alberta, Canada T6G 2R3.

## Abstract

Respiratory viruses are capable of transmitting via an aerosol route. Emerging evidence suggests that SARS-CoV-2 which causes COVID-19 can be spread through airborne transmission, particularly in indoor environments with poor ventilation. Heating, ventilation, and air conditioning (HVAC) systems can play a role in mitigating airborne virus transmission. We conducted a systematic review of the scientific literature examining the effectiveness of HVAC design features in reducing virus transmission—here we report results for ultraviolet (UV) radiation. Following international standards for systematic reviews, we conducted a comprehensive search and synthesized findings from 32 relevant studies published between 1936 and 2020. Research demonstrates that: viruses and bacteriophages are inactivated by UV radiation; increasing UV dose is associated with decreasing survival fraction of viruses and bacteriophages; increasing relative humidity is associated with decreasing susceptibility to UV radiation; UV dose and corresponding survival fraction are affected by airflow pattern, air changes per hour, and UV device location; and UV radiation is associated with decreased transmission in both animal and human studies. This comprehensive synthesis of the scientific evidence examining the impact of UV radiation on virus transmission can be used to guide implementation of systems to mitigate airborne spread and identify priorities for future research.

**Practical Implications:** In-duct ultraviolet germicidal irradiation (UVGI) addresses virus transmission throughout the heating, ventilation, and air conditioning (HVAC) system of a building as a whole; whereas, upper-room UVGI addresses virus transmission in one room of that building. The susceptibility of a virus is often determined from the relationship between UV dose and survival fraction of the virus, and can be affected by relative humidity. Modelling studies revealed that practical implementation of UVGI in HVAC systems should consider airflow patterns, air changes per hour, and UV device location. Future field studies of UVGI systems could address an existing research gap and provide important information on system performance in real-world situations.

## Introduction

COVID-19, the disease caused by a coronavirus (SARS-CoV-2), was declared a pandemic by the World Health Organization in March 2020.^1^ Since then, public health authorities worldwide have sought evidence about the route of transmission and appropriate public health measures to mitigate virus spread. Certain viruses have been proven capable of transmitting via an aerosol route.^2^ In the case of aerosol transmission, virus-laden aerosols are expelled by humans and remain airborne for extended periods of time. Emerging evidence suggests that the SARS-CoV-2 virus can spread through airborne transmission under certain circumstances, particularly in indoor environments with poor ventilation.^3,4^ Selecting appropriate measures to protect the occupants of indoor spaces based on informed, interdisciplinary research is critical to managing the spread of infectious disease.^5^

Heating, ventilation, and air conditioning (HVAC) systems can play a role in mitigating the airborne transmission of viruses by removing or diluting contaminated air inside a building enclosure where humans breathe.^5-7^ Many features within HVAC systems can influence transmission, such as ventilation rates, filters, humidity, and ultraviolet (UV) radiation. Under ultraviolet germicidal irradiation (UVGI), a dose of UV light is delivered to the aerosolized virus which causes damage to the DNA impeding its ability to replicate. The ability for these cells to infect a host are therefore lost.^8^

UV radiation can be applied within mechanically ventilated spaces in the building environment through in-duct or upper-room lamp fixtures. Irradiation in an enclosed space, such as in-duct UVGI, allows for better control of the UV dose, resulting in better control of particle/pathogen exposure to UV radiation. Non-enclosed systems, such as upper-room UVGI, depend on air circulation to drive the particles/pathogens to an irradiated zone near the ceiling (designed to shield unwanted exposure of skin and eyes to UV radiation) as air moves through the UV zone generally due to air currents which are subject to room-scale turbulence. Importantly, in-duct UVGI addresses virus transmission throughout the building by treating the air in the HVAC system; whereas, upper-room UVGI addresses virus transmission within one room by treating the air in that room.

The use of UV radiation as a method of disinfection to help reduce the circulation and transmission of viruses has been investigated in prior research as early as the 1940s. In a narrative review of prevention and control measures of viral bioaerosols, Bing-Yuan^7^ cited several studies that collectively demonstrate the effectiveness of UV in protecting humans from transmission of airborne viruses.^9-13^ A more recent narrative review by Raeiszadeh and Adeli^14^ discussed the use of UV disinfection systems for both surfaces and air in the context of COVID-19, and cites one experimental study demonstrating inactivation of airborne coronaviruses by UV.^15^ Both of these reviews were not systematic and do not provide a comprehensive synthesis of the scientific evidence examining UV radiation.

Even as late as 2019, the American Society of Heating, Refrigerating, and Air-Conditioning Engineers (ASHRAE), in their 2019 ASHRAE Handbook,^16^ recognized that despite improved UVGI system design guidance from significant advances in the analysis and modelling of UVGI systems by Riley et al,^17^ First et al,^18^ Kowalski,^19^ and the National Institute for Occupational Safety and Health (NIOSH),^20^ no consensus guidelines exist that exhaustively address all aspects of UVGI system design.

We conducted a systematic review to examine whether virus transmission is affected by heating, ventilation, and air conditioning (HVAC) design features, in particular, UV radiation. Our objective was to examine published research evaluating the effectiveness of UVGI in reducing virus transmission. The insight drawn from this review could help answer questions of the utility of UVGI as an adjunct technology to curb the spread of the SARS-CoV-2 in mechanically ventilated indoor environments. Further, understanding effectiveness relative to technology set-up and UV dose could inform control measures.

## Methods

This paper describes the results of a systematic review to identify and synthesize the scientific literature examining the impact of UV radiation on virus viability and transmission within the built environment. This was part of a larger research program to review the literature on HVAC design features and airborne virus transmission. Due to the volume and heterogeneity of research, results for other design features of interest (ventilation, filtration, and humidity) are reported separately. We developed an *a priori* protocol^21^ that is publicly available and the systematic review^22^ is registered. We followed standards for the conduct of systematic reviews defined by the international Cochrane organization^23^ with modifications for questions related to etiology.^24^ We report the review according to accepted reporting standards.^25^

### Search strategy

A research librarian (GMT) searched three electronic databases (Ovid MEDLINE, Compendex, Web of Science Core) from inception to June 2020 using concepts related to virus, transmission, and HVAC. The search strategy for Ovid MEDLINE appears in Table 1; the strategies were peer-reviewed by two librarians (TL, AH) prior to implementing the searches. The search was updated in January 2021. We screened reference lists of all relevant papers as well as relevant review articles. We identified conference abstracts through Compendex and Web of Science; abstracts were not included but we searched the literature to see whether any potentially relevant abstracts had been published as complete papers. We did not limit the search by year or language of publication; however, we only included English-language studies due to the volume of available literature and resource constraints. References were managed in EndNote with duplicate records removed prior to screening.

**Table 1:** Search Strategy for Ovid MEDLINE^21^. Database: Ovid MEDLINE(R) ALL 1946 to Present Search Strategy:

| # | Searches |
| --- | --- |
| 1 | exp Aerosols/ |
| 2 | Air Microbiology/ |
| 3 | exp Viruses/ |
| 4 | (aerosol or aerosols or bioaerosol or bioaerosols).mp. |
| 5 | droplet nuclei.mp. |
| 6 | infectio*.mp. |
| 7 | (pathogen or pathogens).mp. |
| 8 | (virus or viruses or viral or virome).mp. |
| 9 | or/1-8 [MeSH + Keywords – Virus concept] |
| 10 | Air Conditioning/ |
| 11 | Air Filters/ or Filtration/ |
| 12 | Humidity/ |
| 13 | Ventilation/ |
| 14 | Ultraviolet Rays/ |
| 15 | air condition*.mp. |
| 16 | (air change rate or air change rates or air changes per hour or air exchange rate or air exchange rates or air exchanges per hour).mp. |
| 17 | (airflow or air flow).mp. |
| 18 | built environment.mp. |
| 19 | computational fluid dynamics.mp. |
| 20 | ((distance adj6 index) or long distances).mp. |
| 21 | HVAC.mp. |
| 22 | (filter or filters or filtration).mp. |
| 23 | humidity.mp. |
| 24 | (ultraviolet or UV).mp. |
| 25 | ventilat*.mp. |
| 26 | or/10-25 [MeSH + Keywords – HVAC concept] |
| 27 | Air Pollution, Indoor/ |
| 28 | exp Disease Transmission, Infectious/ |
| 29 | (indoor adj1 (air quality or environment*)).mp. |
| 30 | transmission.mp. |
| 31 | or/27-30 [MeSH + Keywords – Transmission concept] |
| 32 | 9 and 26 and 31 |
| 33 | remove duplicates from 32 |

MeSH = Medical Subject Headings

### Study selection

Study selection occurred in two stages. First, two reviewers independently screened the titles and abstracts of all references identified by the electronic databases searches. Relevance of each record was classified as Yes, No or Maybe. Conflicts between Yes/Maybe and No were resolved by one reviewer. We conducted pilot testing with three sets of studies (n=199 each) to ensure consistency among the review team. After each set of pilot screening, the review team met to discuss discrepancies and develop decision rules. The second stage involved two reviewers independently reviewing the full text articles and applying the inclusion/exclusion criteria. Studies were classified as Include or Exclude. Conflicts between Include and Exclude were resolved by consensus of the review team. Conflicts between different exclusion reasons were resolved by one reviewer. We pilot tested the second stage of screening with three sets of studies (n=30 each). After each pilot round, the review team met to resolve discrepancies. We conducted screening using Covidence software.

### Inclusion and exclusion criteria

Table 2 lists our inclusion and exclusion criteria. As noted above, this systematic review was part of a larger effort to examine different HVAC design features and virus transmission. We searched and screened for all design features at once, but only studies evaluating UV radiation are synthesized here. While our interest was UV within HVAC systems, we also included studies of upper room UVGI because of its similar utility and mechanism of air disinfection. We searched for a variety of agents but prioritized studies of viruses or agents that simulated viruses; we planned to include other agents (e.g., bacteria, fungi) only if studies were not available that were specific to viruses. We included studies of bacteriophages, which are viruses that infect bacterial cells.^19^ We were interested in studies of the indoor built environment (e.g., office, public, residential buildings) which had mechanical ventilation. We included primary research that provided quantitative results of the correlation or association between installed UV radiation and virus survival or transmission. We placed no restrictions on year of publication; we included only English-language, peer-reviewed publications.

**Table 2.** Inclusion and exclusion criteria for systematic review^21^.

| Item | Inclusion criteria | Exclusion criteria |
| --- | --- | --- |
| Agent | • Viruses<br>• Aerosols<br>• Bioaerosols<br>• Droplet nuclei<br>• Other pathogens (e.g., bacteria, fungi)<br>*We planned a staged process: if we identified studies specific to viruses for each HVAC design feature, we would not include other pathogens; however, for design features where we did not find studies specific to viruses, we would expand to other pathogens.* |  |
| HVAC | Design features relating to:<br>• Ventilation (ventilation rate, air changes per hour (ACH), air exchange, airflow pattern, pressurization)<br>• Filtration (air filtration, filter type, MERV rating, filter age and/or use, pressure drop, holding capacity, replacement, change frequency)<br>• Ultraviolet germicidal irradiation (UVGI; power, dose, uniformity of dose, flow rate, bioaerosol inactivation efficiency, location)<br>• Humidity or relative humidity | Examines HVAC / mechanical / or other ventilation mechanisms overall, but not by specific design features. |
| Setting | • Office buildings<br>• Public buildings (e.g., schools, day cares)<br>• Residential buildings<br>• Hospitals and other healthcare facilities (e.g., clinics)<br>• Transport vehicles (e.g., aircraft) or hubs (e.g., airports) | • Outdoor settings<br>• Indoor settings with natural ventilation |
| Outcomes | Quantitative data evaluating the correlation or association between virus transmission and above HVAC features | Qualitative data |
| Study design | Primary research, including:<br>• Epidemiological studies<br>• Observational studies (e.g., cohort, case-control, cross-sectional)<br>• Experimental studies (including human or animal)<br>• Modelling studies, including CFD | • Review articles<br>• Commentaries, opinion pieces<br>• Qualitative studies |
| Language | English<br>*We planned a staged process where we would include studies in languages other than English if we do not identify English language studies for specific HVAC design features or if we identified clusters of potentially relevant studies in another language.* |  |
| Year | No restrictions |  |
| Publication status | Published, peer-reviewed | Unpublished, not peer-reviewed |

CFD = computational fluid dynamics; HVAC = heating, ventilation, and air conditioning; MERV = minimum efficiency reporting value; UVGI = ultraviolet germicidal irradiation

### Risk of Bias assessment

For experimental studies, we assessed risk of bias based on three key domains: selection bias, information bias and confounding.^26-27^ We assessed each domain as high, unclear, or low risk of bias using signaling questions^28^ from guidance documents for the different study types we included; e.g., animal studies, laboratory experiments, epidemiological studies.^26-27,29^ For modelling studies, we assessed the following three key domains: definition, assumption, and validation.^29-30^ Definition considers model complexity and data sources, assumption considers the description and explanation of model assumptions, and validation considered model validation and sensitivity analysis.^30^ Also, we assessed each domain as high, unclear or low risk of bias based on signaling questions.^29-31^ The risk of bias items were pilot tested among three review authors, then two reviewers (GMT, BAF) applied the criteria independently to each relevant study and met to resolve discrepancies.

### Data extraction

We extracted general information about the study (authors, year of publication, country of corresponding author, study design) and methods (setting, population [as applicable], agent studied, intervention set-up). We extracted details on UV treatment parameters (where available), including: wavelength; UV dose; exposure time; and fluence rate. Also, we extracted information (where available) regarding relative humidity (RH). The studies were grouped as “in-duct UVGI” and “upper-room UVGI.” We extracted quantitative data, as well as results of any tests of statistical significance related to UV features. A priori, our primary outcome of interest was quantitative measures of the association between UV radiation and virus transmission; however, during the review we realized that most studies focused on proxy variables such as virus survival. Therefore, we extracted data on actual transmission where available (i.e., infections), as well as proxy variables (e.g., survival fraction (SF), dose-response of UV dose and survival fraction, susceptibility (Z), and equivalent air changes per hour (ACH) due to UV radiation (ACHuv)). Survival fraction (SF) is the concentration of virus after UV exposure divided by the concentration of virus before UV exposure. UV dose (*D*) [J/m^2^] is the fluence rate [W/m^2^] multiplied by the exposure time [s]. The dose-response relationship of UV dose and survival fraction is often represented as *SF* = exp(-*ZD*), where *Z* is the susceptibility and exp() represents exponential function. Equivalent ACH due to UV radiation (ACHuv) is the number of air changes per hour (ACH) that would produce the same reduction in virus concentration as obtained using UV radiation. We created a data extraction form spreadsheet to ensure comprehensive and consistent capture of data. One reviewer extracted data and a second reviewer verified data for accuracy and completeness. Discrepancies were discussed by the review team.

### Data synthesis

We anticipated that meta-analysis would not be possible due to heterogeneity across studies in terms of study design, UV features examined, outcomes assessed, and reporting of results. We developed evidence tables describing the studies and their results (as reported by the authors of the primary studies). We provide a narrative synthesis of the results of relevant studies. To allow for meaningful synthesis and comparison across studies, we divided the studies into four groups: aerosolized virus, modelling, animal studies, human studies. Within the aerosolized virus group, the effect of RH was further examined.

## Results

The electronic searches and other sources yielded 12,177 unique citations; 2,428 were identified as potentially relevant based on title/abstract screening and 568 met the review’s inclusion criteria (Figure 1). Of the 568, 125 were relevant to UV radiation and, of those 125, 32 were relevant to UV radiation and virus (Figure 1). Among the 32 relevant studies there were: 16 aerosolized virus and bacteriophage studies (Table 3), 7 modelling studies (Table 4), 4 animal studies (Table 5), and 5 human studies (Table 6). Studies were published between 1936 and 2020 (median year 2007.5). While the majority of the experimental and modelling studies were published between 2005 and 2020, with one exception in 1964, the human studies are all from the 1940s and the animal studies spanned from 1936 to 2020. The majority of studies were conducted in the United States (n=24). Studies were funded by national research funding organizations (n=13), industry (n=6), a university and state grant (n=1), and hospital (n=1); 2 studies reported no external funding and 8 studies did not report funding source.

**Figure 1.**
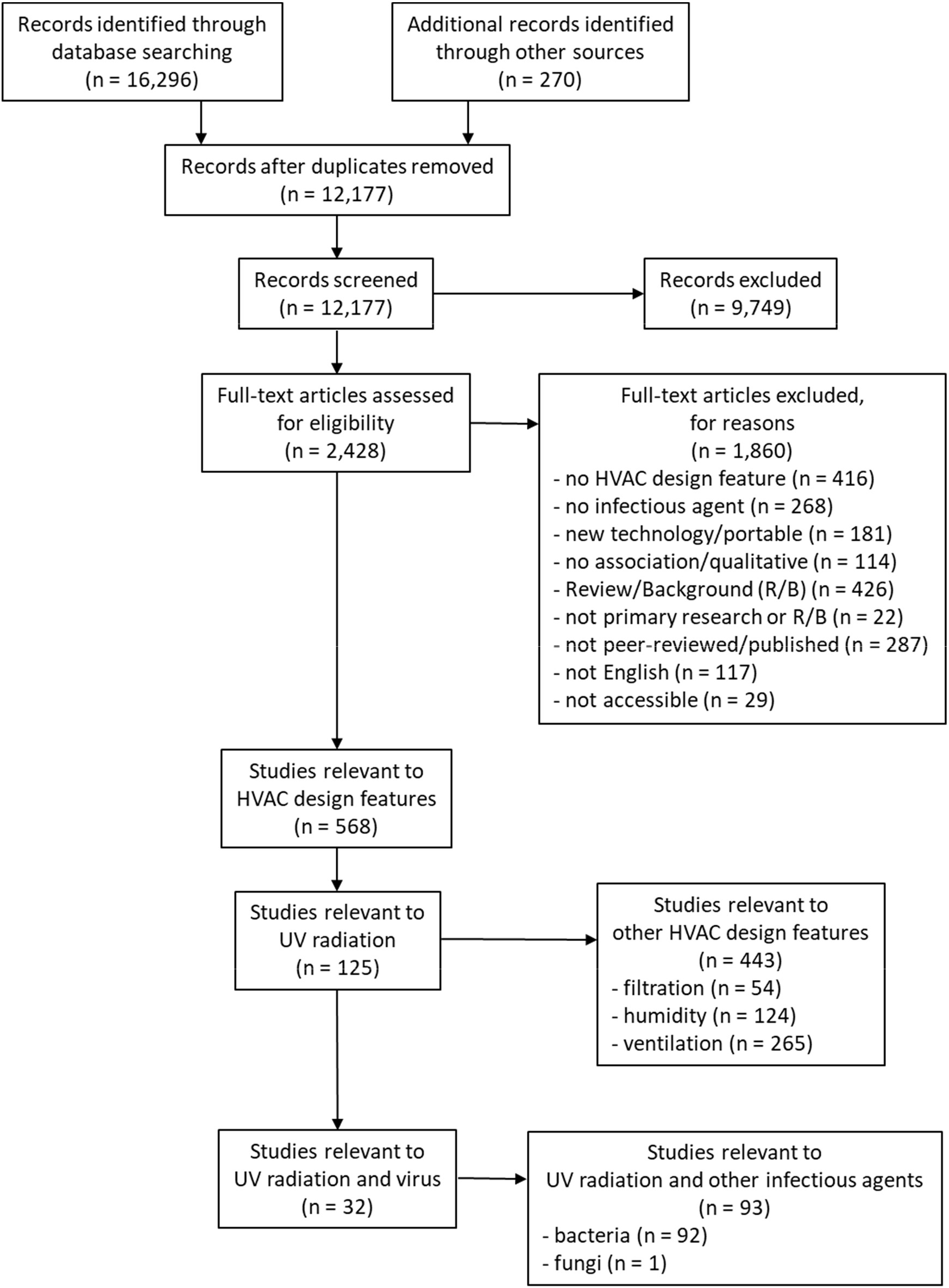
Flow of studies through the selection process (note: search was conducted for all HVAC design features but only studies of UV radiation are included in this manuscript).

**Table 3.** Summary of characteristics and findings for aerosolized virus and bacteriophage studies of UV treatments.

| Author<br>(Year)<br>Country | Infectious<br>Agent | Treatment | Outcome<br>Parameter | Data | Association |
| --- | --- | --- | --- | --- | --- |
| Jensen<br>(1964)<sup>36</sup><br>USA | Influenza A<br>(WSN)<br>Vaccinia virus<br>Adenovirus<br>(type 2)<br>Coxsackie B1<br>Sindbis | Wavelength:<br>253.7 nm<br>Dose:<br>> 19.4 J/m<sup>2</sup><br>Exposure time:<br>0.3 s, 0.6 s<br>RH:<br>Influenza A at<br>68%RH,<br>Vaccinia virus<br>at 65%RH,<br>Adenovirus at<br>50%RH,<br>Coxsackie B1<br>at 66%RH,<br>Sindbis at<br>62%RH<br>In-duct UVGI | Survival fraction<br>(SF) from<br>efficiency | Influenza A (at 68%RH)<br>SF= 0.0014 at 0.3 s<br>SF = 0.0010 at 0.6 s<br>Vaccinia virus (at 65%RH)<br>SF= 0.0004 at 0.3 s<br>SF = 0.0001 at 0.6 s<br>Adenovirus (at 50%RH)<br>SF= 0.0869 at 0.3 s<br>SF = 0.0312 at 0.6 s<br>Coxsackie B1 (at 66%RH)<br>SF= 0.0240 at 0.3 s<br>SF = 0.0005 at 0.6 s<br>Sindbis (at 62%RH)<br>SF= 0.0327 at 0.3 s<br>SF = 0.0047 at 0.6 s | - Increasing<br>exposure time<br>(related to<br>increasing dose)<br>associated with<br>decreasing<br>survival fraction. |
| Tseng<br>(2005)<sup>32</sup><br>Taiwan | MS2 (ssRNA)<br>[15597-B1]<br>phiX174<br>(ssDNA)<br>phi6 (dsRNA,<br>enveloped)<br>T7 (dsDNA) | Wavelength:<br>253.7 nm<br>Dose: < 12 J/m<sup>2</sup><br>(Figure 2 and 3,<br>p.1140)<br>RH: 55%RH;<br>85%RH<br>Chamber:<br>cylinder<br>In-duct UVGI | Dose-response<br>Susceptibility (Z)<br>*Effect of RH* | MS2 (ssRNA)<br>Z=0.81 m<sup>2</sup>/J at 55%RH<br>Z=0.64 m<sup>2</sup>/J at 85%RH<br>phiX174 (ssDNA)<br>Z=0.71 m<sup>2</sup>/J at 55%RH<br>Z=0.53 m<sup>2</sup>/J at 85%RH<br>phi6 (dsRNA)<br>Z=0.43 m<sup>2</sup>/J at 55%RH<br>Z=0.31 m<sup>2</sup>/J at 85%RH<br>T7 (dsDNA)<br>Z=0.33 m<sup>2</sup>/J at 55%RH<br>Z=0.22 m<sup>2</sup>/J at 85%RH<br>-Z significantly lower at<br>higher RH for all four<br>bacteriophages.<br>- ssRNA and ssDNA had<br>increased susceptibility<br>compared with dsRNA and<br>dsDNA.<br>- ssRNA and ssDNA had<br>greater increased<br>susceptibility with increased<br>RH compared with dsRNA<br>and dsDNA. | - Increasing dose<br>associated with<br>decreasing<br>survival fraction.<br>- Increasing RH<br>associated with<br>decreasing<br>susceptibility. |
| Walker<br>(2007)<sup>10</sup><br>USA | Murine<br>hepatitis virus<br>(MHV)<br>coronavirus<br>- enveloped | Wavelength:<br>254 nm<br>Dose: 5.99 J/m<sup>2</sup><br>Exposure time:<br>16.2 s<br>RH: 50%RH | Survival fraction<br>(SF)<br>Susceptibility (Z)<br>(Susceptibility<br>calculated from<br>survival fraction, | At 50%RH,<br>SF = 0.122 ± 0.072<br>Z = 0.377 ± 0.119 m<sup>2</sup>/J | - UV radiation<br>associated with<br>survival fraction. |
|  | (p.5463) | Chamber:<br>experimental<br>duct<br>In-duct UVGI | not dose-response.) | - Coronavirus had increased<br>susceptibility compared with<br>MS2 and adenovirus. |  |
| | MS2 [15597-B1]<br>- not<br>enveloped<br>(p.5463) | Wavelength:<br>254 nm<br>Dose:<br>26.08 J/m<sup>2</sup><br>Exposure time:<br>16.2 s<br>RH:<br>32%-50%RH,<br>74%-82%RH<br>Chamber:<br>experimental<br>duct<br>In-duct UVGI | Survival fraction<br>(SF)<br>Susceptibility (Z)<br>(Susceptibility<br>calculated from<br>survival fraction,<br>not dose-response.)<br>*Effect of RH* | At 32%-50%RH,<br>SF = $0.311 \pm 0.029$<br>Z = $0.038 \pm 0.003 \text{ m}^2/\text{J}$<br>At 74%-82%RH,<br>SF = $0.246 \pm 0.035$<br>Z = $0.048 \pm 0.005 \text{ m}^2/\text{J}$ | - Increasing RH<br>associated with<br>increasing<br>susceptibility. |
| | Adenovirus<br>(serotype 2)<br>- dsDNA<br>(p.5463) | Wavelength:<br>254 nm<br>Dose:<br>26.08 J/m<sup>2</sup><br>Exposure time:<br>16.2 s<br>RH:<br>27%-40%RH,<br>50%-55%RH,<br>76%-80%RH<br>Chamber:<br>experimental<br>duct<br>In-duct UVGI | Survival fraction<br>(SF)<br>Susceptibility (Z)<br>(Susceptibility<br>calculated from<br>survival fraction,<br>not dose-response.)<br>*Effect of RH* | At 27%-40%RH,<br>SF = $0.329 \pm 0.023$<br>Z = $0.038 \pm 0.003 \text{ m}^2/\text{J}$<br>At 50%-55%RH,<br>SF = $0.206 \pm 0.035$<br>Z = $0.052 \pm 0.004 \text{ m}^2/\text{J}$<br>At 76%-80%RH,<br>SF = $0.136 \pm 0.005$<br>Z = $0.068 \pm 0.002 \text{ m}^2/\text{J}$ | - Increasing RH<br>associated with<br>increasing<br>susceptibility. |
| McDevitt<br>(2007)<sup>11</sup><br>USA | Vaccinia virus<br>(strain WR)<br>- enveloped<br>- dsDNA<br>(p.5763) | Wavelength:<br>254 nm<br>Dose: 0.1 -<br>3.2 J/m<sup>2</sup><br>Exposure time:<br>7.6 s<br>RH:<br>18%-23%RH,<br>58%-63%RH,<br>78%-83%RH<br>Chamber:<br>Benchtop<br>In-duct UVGI | Dose-response<br>Susceptibility (Z)<br>*Effect of RH* | In SRF,<br>at 18%-23%RH,<br>Z=6.16 (4.27-8.89) m<sup>2</sup>/J;<br>at 58%-63%RH,<br>Z=1.94 (1.66-2.26) m<sup>2</sup>/J;<br>At 78%-83%RH<br>Z=1.63 (1.14-2.32) m<sup>2</sup>/J;<br>In water,<br>at 18%-23%RH,<br>Z=9.48 (5.32-16.90) m<sup>2</sup>/J;<br>at 58%-63%RH,<br>Z=2.54 (2.05-3.16) m<sup>2</sup>/J;<br>at 78%-83%RH<br>Z=1.42 (1.15-1.75) m<sup>2</sup>/J<br>-Z significantly lower at<br>higher RH after controlling<br>for medium.<br>-Medium significant overall<br>after controlling for RH. | - Increasing dose<br>associated with<br>decreasing<br>survival fraction.<br>- Increasing RH<br>associated with<br>decreasing<br>susceptibility. |
| Su<br>(2007)<sup>58</sup><br>USA | Fluorescent<br>bioaerosols | Safety<br>requirement for<br>occupants:<br>0.002 W/m<sup>2</sup> | Fluorescent<br>bioaerosol counts<br>(FBC) | For 20 days evaluated,<br>12 days had significantly<br>lower FBC in UVGI rooms<br>compared with non-UVGI<br>rooms, 6 days had | - UV radiation<br>associated with<br>reduction of<br>fluorescent<br>bioaerosol counts |
|  |  | Setting: Public elementary school<br>Upper-room UVGI |  | *significantly* greater FBC in UVGI rooms than non-UVGI rooms, and 2 days were *not statistically different* between UVGI and non-UVGI rooms. | (FBC) on 12 days of the 20 days evaluated. |
| First (2007)<sup>41</sup>; Rudnick (2007)<sup>42</sup> USA | Vaccinia virus (Western Reserve strain) | Wavelength: 254 nm<br>Mean Room Fluence Rate (Rudnick, 2007<sup>42</sup>)<br>0.0177 W/m<sup>2</sup>; 0.140 W/m<sup>2</sup><br>RH: 50%RH<br>Chamber: room<br>Upper-room UVGI | Survival fraction (SF)<br>Equivalent ACH (ACHuv)<br>Susceptibility (Z) | At 0.0177 W/m<sup>2</sup>,<br>SF = 0.10 ± 0.05<br>ACHuv = 19.0 ACH<br>At 0.140 W/m<sup>2</sup>,<br>SF = 0.04 ± 0.02<br>ACHuv = 42.8 ACH<br>Z = 1.0 m<sup>2</sup>/J<br>[First (2007)<sup>41</sup> p.325; Rudnick (2007)<sup>42</sup> p.356, p.362] | - Increasing fluence rate (related to increasing dose) associated with decreasing survival fraction and increasing equivalent ACH. |
| McDevitt (2008)<sup>12</sup> USA | Vaccinia virus (Western Reserve strain) | Wavelength: 254 nm<br>Number of fixtures: 1, 4<br>ACH: 2, 6 ACH<br>Condition: Winter (40%RH, fan upwards); Summer (80%RH, fan downwards)<br>Chamber: room<br>[Note: same chamber as First (2007) and Rudnick (2007)]<br>[For 6 ACH and 1 fixture, dose was 17 J/m<sup>2</sup> and was expected to be 4-times higher with 4 fixtures (p.5)].<br>Upper-room UVGI | Survival fraction (SF)<br>Equivalent ACH (ACHuv) | For Winter, 2 ACH and 1 fixture<br>SF=0.017 (0.014-0.021)<br>ACHuv=110 (93-140) ACH<br>For Winter, 2 ACH and 4 fixtures<br>SF=0.003 (0.002-0.005)<br>ACHuv=580 (410-830) ACH<br>For Winter, 6 ACH and 1 fixture<br>SF=0.038 (0.032-0.046)<br>ACHuv=150 (120-180) ACH<br>For Winter, 6 ACH and 4 fixtures<br>SF=0.006 (0.004-0.008)<br>ACHuv=1000 (740-1400) ACH<br>For Summer, 2 ACH and 1 fixture<br>SF=0.087 (0.062-0.120)<br>ACHuv=18 (15-30) ACH<br>For Summer, 2 ACH and 4 fixtures<br>SF=0.061 (0.053-0.071)<br>ACHuv=31 (26-36) ACH<br>For Summer, 6 ACH and 1 fixture<br>SF=0.140 (0.120-0.160)<br>ACHuv=38 (31-46) ACH<br>For Summer, 6 ACH and 4 fixtures<br>SF=0.078 (0.065-0.084)<br>ACHuv=71 (58-86) ACH | - Increasing number of fixtures (related to increasing dose) associated with decreasing survival fraction and increasing equivalent ACH.<br>- Increasing ACH associated with increasing survival fraction and increasing equivalent ACH.<br>- Comparing Winter and Summer, increasing RH and changing fan direction associated with increasing survival fraction and decreasing equivalent ACH. |
| Terrier (2009)<sup>37</sup> France | Influenza A (H5N2) [A/Finch/Engl and/2051/2021 (H2N5)] | Wavelength: 254 nm<br>Chamber: | Survival Fraction (SF) from efficiency | Influenza A (H2N5)<br>SF = 0.0040<br>hPIV-3<br>SF = 0.0003<br>RSV | - UV radiation associated with survival fraction. |
|  | human parainfluenza 3 (hPIV-3)<br>Respiratory syncytial virus (RSV) | experimental duct<br>In-duct UVGI |  | SF = 0.0080 |  |
| McDevitt (2012)<sup>13</sup><br>USA | Influenza A (H1N1)<br>[A/PR/8/34 H1N1]<br>- RNA (p.1668) | Wavelength: 254 nm<br>Dose: 4.9 - 15 J/m<sup>2</sup><br>RH: 25%-27%RH, 50%-54%RH, 81%-84%RH<br>Chamber: benchtop<br>In-duct UVGI | Dose-response Susceptibility (Z)<br>*Effect of RH* | At 25%-27%RH, Z=0.29 (0.27-0.31) m<sup>2</sup>/J<br>At 50%-54%RH, Z = 0.27 (0.26-0.31) m<sup>2</sup>/J<br>At 81%-84%RH, Z = 0.22 (0.21-0.23) m<sup>2</sup>/J<br>- Z *significantly* lower at higher RH | - Increasing dose associated with decreasing fraction surviving.<br>- Increasing RH associated with decreasing susceptibility. |
| Cutler (2012)<sup>33</sup><br>USA | Porcine Reproductive and Respiratory Syndrome Virus (PRRSV) | Wavelength: 254 nm<br>Dose: 0, 0.5, 1.2, 2 J/m<sup>2</sup><br>Exposure time: 0, 0.07, 0.14, 0.25 s<br>RH: ≤24%RH, 25%-79%RH, ≥80%RH<br>(Note: large range of RH for 25%-79%RH)<br>Chamber: two reservoirs<br>In-duct UVGI | Dose-response Susceptibility (Z)<br>*Effect of RH* | At ≤24%RH, Z=0.425 m<sup>2</sup>/J<br>At, 25-79%RH Z=0.587 m<sup>2</sup>/J<br>At ≥80%RH, Z=0.341 m<sup>2</sup>/J<br>- Z *significantly* lower at ≥80%RH compared with 25%-79%RH. | -Increasing dose associated with decreasing survival fraction.<br>- Increasing RH associated with decreasing susceptibility comparing 25%-79%RH with ≥80%RH. |
| Verreault (2015)<sup>40</sup><br>Canada | MS2 (ssRNA) [15597-B1]<br>phiX174 (ssDNA)<br>phi6 (dsRNA, enveloped)<br>PR772 (dsDNA) | Wavelength: 254 nm<br>Dose: UV sensor data not reported<br>Exposure time: 3 s, 6 s, 10 s<br>RH: 20%RH<br>Chamber: rotating drum | Relative Infectious Ratio<br>(Note: relative infectious ratio appears to be equivalent to survival fraction) | - Relative infectious ratio *significantly* lower at higher exposure time.<br>- MS2 (ssRNA) greater relative infectious ratio compared with other bacteriophages. | - Increasing exposure time associated with decreasing relative infectious ratio. |
| Lin (2017)<sup>35</sup><br>Canada | phi6<br>- RNA (p.555) | Pulsed UVGI<br>Wavelength: 200-280 nm<br>Cumulative dose: 14, 28, 43 J/m<sup>2</sup><br>RH: 41%-58%RH<br>Chamber: cubic chamber with 1.8 m sides<br>Upper-room UVGI | “fast decay” Survival fraction (SF)<br>Susceptibility (Z) (Susceptibility calculated from survival fraction, not dose-response.)<br>“slow decay” Dose-response Susceptibility (Z) | “fast decay” SF=0.035 ± 0.024<br>Z=0.24 m<sup>2</sup>/J<br>“slow decay” Z=0.02 m<sup>2</sup>/J | - Increasing dose associated with decreasing survival fraction. |
| Welch (2018)<sup>34</sup><br>USA | Influenza A (H1N1)<br>[A/PR/8/34 {H1N1}] | Wavelength: 222 nm<br>Dose: 0, 8, 13, 20 J/m<sup>2</sup><br>RH: 55%RH<br>Chamber: Benchtop<br>In-duct UVGI | Dose-response Susceptibility (Z) | Z=0.18 (0.15-0.21) m<sup>2</sup>/J | - Increasing dose associated with decreasing survival fraction. |
| Buonanno (2020)<sup>15</sup><br>USA | Coronavirus 229E<br>Coronavirus OC43<br>229E -alpha (p.2)<br>OC43 -beta (p.2)<br>SARS-CoV-2 - beta (p.2) | Wavelength: 222 nm<br>Dose: 0, 5, 10, 20 J/m<sup>2</sup><br>RH: 66%RH<br>Chamber: Benchtop<br>In-duct UVGI | Dose-response Susceptibility (Z) | Coronavirus 229E Z=0.41 (0.25-0.48) m<sup>2</sup>/J<br>Coronavirus OC43 Z=0.59 (0.38-0.71) m<sup>2</sup>/J | - Increasing dose associated with decreasing survival fraction. |
| Pearce-Walker (2020)<sup>39</sup><br>USA | MS2 [15579-B]<br>canine distemper virus (CDV) | Wavelength: 253.7 nm<br>2 sets of 2 lamps @ 0.6 W/m<sup>2</sup><br>RH: 12%-50%RH<br>Chamber: HVAC duct<br>In-duct UVGI | Survival fraction (SF) from log reduction<br>(For CDV, log reduction calculated using lower detection limit = 3.16 TCID<sub>50</sub>/mL) | MS2 SF=0.003<br>CDV SF<0.03 | - UV radiation associated with survival fraction. |
| Qiao (2020)<sup>38</sup><br>USA | Porcine respiratory coronavirus [VR-2384]<br>-alpha (p.B; p.E) | Wavelength: 252.7±1 nm<br>Dose: 139.2, 202.8, 496.3 J/m<sup>2</sup><br>Exposure time: 1.25, 1.81, 4.44 s<br>RH: 57%-62%RH<br>Chamber: wind tunnel<br>In-duct UVGI | Survival fraction (SF) from efficiency<br>(Efficiency calculated using lower detection limit = 3.16 x10<sup>1</sup> TCID<sub>50</sub>/mL) | SF<0.0060 at 139.2 J/m<sup>2</sup><br>SF<0.0004 at 202.8 J/m<sup>2</sup><br>SF<0.0002 at 496.3 J/m<sup>2</sup> | - UV radiation associated with survival fraction. |

Data reported as (95% confidence interval) or ± standard deviation.
ssRNA = single-stranded ribonucleic acid; ssDNA = single-stranded deoxyribonucleic acid;
dsRNA = double-stranded ribonucleic acid; dsDNA = double-stranded deoxyribonucleic acid

**Table 4.** Summary of characteristics and findings for modelling studies of UV treatments.

| Author<br>(Year)<br>Country | Infectious<br>Agent | Model | Outcome<br>Parameter | Data | Association |
| --- | --- | --- | --- | --- | --- |
| Noakes<br>(2006)<sup>47</sup><br>UK | Infectious<br>agents<br>sensitive to<br>UV radiation<br>including<br>viruses | UV device and<br>ventilation<br>configuration<br>Setting: single<br>patient hospital<br>room<br>CFD model<br>Upper-room UVGI | UV dose<br>distribution<br>Volume average<br>UV dose where<br>volume could be<br>room, upper-room or lower-<br>room | - Higher average UV<br>dose in the occupied<br>region of the room<br>when ventilated air<br>supplied at floor and<br>extracted at ceiling<br>compared with<br>supplied at ceiling and<br>extracted at floor.<br>- UV dose less affected<br>by UV device location<br>when ventilated air<br>supplied at floor and<br>extracted at ceiling<br>compared with<br>supplied at ceiling and<br>extracted at floor. | - UV dose affected by<br>airflow pattern.<br>- UV dose affected by<br>UV device location. |
| First<br>(2007)<sup>41</sup>;<br>Rudnick<br>(2007)<sup>42</sup><br>USA | Vaccinia virus<br>(Western<br>Reserve<br>strain) | Wavelength:<br>254 nm<br>Mean Room<br>Fluence Rate<br>(Rudnick, 2007<sup>42</sup>)<br>0.0177 W/m<sup>2</sup>;<br>0.140 W/m<sup>2</sup><br>RH: 50%RH<br>Two box model<br>Upper-room UVGI | Survival fraction<br>(SF)<br>Effectiveness<br>index (EI) which<br>considers vertical<br>mixing and UV<br>radiation | At 0.0177 W/m<sup>2</sup>,<br>SF = 0.10<br>EI = 18.2<br>At 0.140 W/m<sup>2</sup>,<br>SF = 0.04<br>EI = 36.4 | - Survival fraction (SF)<br>proportional to<br>Effectiveness index<br>(EI) to the -0.74<br>power.<br>- Increasing fluence<br>rate (related to<br>increasing dose)<br>associated with<br>decreasing survival<br>fraction and increasing<br>EI. |
| Li<br>(2010)<sup>43</sup><br>South<br>Korea | Infectious<br>agents<br>sensitive to<br>UV radiation<br>including<br>viruses | UV device<br>configuration: wall,<br>corner<br>Fluence rate: 0, 59,<br>118, 236 W/m<sup>2</sup><br>ventilation: 0, 3,<br>6 ACH<br>Evaluated<br>Z=0.059 m<sup>2</sup>/J<br>(p.51)<br>Setting: hospital<br>isolation room<br>CFD model<br>Upper-room UVGI | Survival fraction<br>(SF)<br>from efficiency<br>(removal SF<br>equals<br>UV radiation SF<br>plus<br>ventilation SF) | -Removal SF lower<br>when UV on wall<br>compared with corner<br>-Removal SF lower<br>with increasing fluence<br>rate<br>- Removal SF lower<br>with increasing ACH<br>- Ventilation SF lower<br>with increasing ACH<br>- UV radiation SF<br>greater with increasing<br>ACH | - UV dose and survival<br>fraction affected by<br>UV device location.<br>- Increasing fluence<br>rate (related to<br>increasing dose)<br>associated with<br>decreasing removal<br>survival fraction.<br>- Increasing ACH<br>associated with<br>increasing UV<br>radiation survival<br>fraction.<br>- Increasing ACH<br>associated with<br>decreasing removal<br>survival fraction and<br>decreasing ventilation<br>survival fraction. |
| Sung (2011)<sup>45</sup><br>Japan | Influenza | UV device configuration<br>Evaluated $Z=0.27 \text{ m}^2/\text{J}$ for influenza A (p.2328)<br>[see McDevitt (2012)<sup>13</sup>]<br>Setting: four patient hospital room<br>CFD model<br>Upper-room UVGI | UV dose distribution<br>Survival fraction (SF) | - In breathing zone of neighbouring patient, SF = 0.26 - 0.52<br>- Highest UV dose and lowest survival fraction when upper-room UVGI installed on opposite side of the one exhaust opening. | UV dose affected by UV device location. |
| Zheng (2016)<sup>48</sup><br>USA/<br>China | Influenza | UVGI equivalent air changes per hour ACH<sub>uv</sub> = 12 ACH<br>SIER epidemic model and contact network model<br>UVGI on cruise ship | Attack rate = Number of new cases in population at risk divided by number of persons at risk in population | Attack rate decreased 87.8% with UVGI compared with baseline (no UV).: 4.08% compared with 33.42% | UV radiation associated with decreased attack rate (related to number of cases). |
| Firrantello (2018)<sup>46</sup><br>USA | Rhinovirus | Coil face: $2 \text{ W}/\text{m}^2$<br>Allowable minimum: $0.50 \text{ W}/\text{m}^2$<br>Evaluated $Z=0.02996 \text{ m}^2/\text{J}$ for virus (p.604)<br>Parametric model of energy, indoor air quality (IAQ), economic benefits<br>UVGI of cooling coil (in-duct UVGI) | IAQ benefit: Work Loss Days (WLD); Hospital Acquired Infections (HAI); Disability Adjusted Life Years (DALY) | “The estimated monetary IAQ benefit from collateral air treatment of a UVGI coil irradiation system treatment was much greater than the estimated energy cost savings.” (p. 609) | UV radiation of coils associated with monetary indoor air quality benefit related to absence, infection, or disability. |
| Buchan (2020)<sup>44</sup><br>UK | Coronavirus | Wavelength: 222 nm<br>Evaluated $Z=0.41 \text{ m}^2/\text{J}$ for coronavirus (Buonanno, 2020<sup>15</sup>) (p.2)<br>Setting: single patient hospital room<br>Coupled radiation-CFD model<br>Upper-room UVGI | Survival fraction (SF) | At 0.8 ACH, SF = 0.15<br>At 8 ACH, SF = 0.43 | Increasing ACH associated with increasing survival fraction. |

CFD = computational fluid dynamics
SEIR = susceptible-exposed-infected-recovered

**Table 5.** Summary of characteristics and findings for animal studies of UV treatments.

| Author (Year) Country | Infectious Agent | Treatment | Outcome Parameter | Data | Association |
| --- | --- | --- | --- | --- | --- |
| Wells (1936)<sup>49</sup><br>USA | Influenza [Puerto Rico 8] | Dose: UV light intensity previous “marked bactericidal effect” (p.412)<br>Setting: virus aerosolizing and animal exposure chamber; treatment tube<br>In-duct UVGI | Transmission<br>Air from UV group and non-UV group inoculated intranasally. | Transmission in 0 of 2 ferrets in UV group and 2 of 2 ferrets in non-UV group. | UV radiation associated with decreased transmission. |
| Jakab (1982)<sup>50</sup><br>USA | Influenza A [Mouse-adapted influenza A/PR8/34] | Dose: 4.2, 8.4, 12.6 J/m<sup>2</sup> *<br>Setting: virus aerosolizing chamber; treatment slot; animal exposure chamber<br>In-duct UVGI | Transmission | 9% mortality of mice in highest dose UV group compared with 100% mortality of mice in non-UV group. | - UV radiation associated with decreased mortality.<br>- Increasing UV dose associated with decreasing mortality. |
| Dee (2006)<sup>52</sup><br>USA | Porcine Reproductive and Respiratory Syndrome Virus (PRRSV) | Wavelength: 253.7 nm<br>Setting: virus aerosolizing chamber; treatment duct; animal exposure chamber<br>In-duct UVGI | Transmission | Transmission in 8 of 10 pigs in UV group *not statistically different* than transmission in 9 of 10 pigs in non-UV group likely due to insufficient exposure time (p.32). | UV radiation did not have a statistically significant effect on transmission likely due to insufficient exposure time (related to UV dose). |
| Jaynes (2020)<sup>51</sup><br>USA | Upper respiratory tract infections (URI) | Dose: designed to eliminate 99% of influenza, feline calicivirus, and bacteria *B bronchiseptica* (p.930)<br>Setting: Kitten nursery<br>Upper-room UVGI | Incidence of URI | Incidence of URI *significantly* decreased 87.1% with UVGI in 2018 compared with no UVGI in 2016: 1.6 cases per 100 kitten admissions compared with 12.4 cases per 100 kitten admissions. | UV radiation associated with decreased incidence of upper respiratory tract infections. |

\* inconsistencies in units for dose appear to be typos in the original paper, we assumed these to be 420, 840 and 1260 $\mu\text{J}/\text{cm}^2$

**Table 6.**
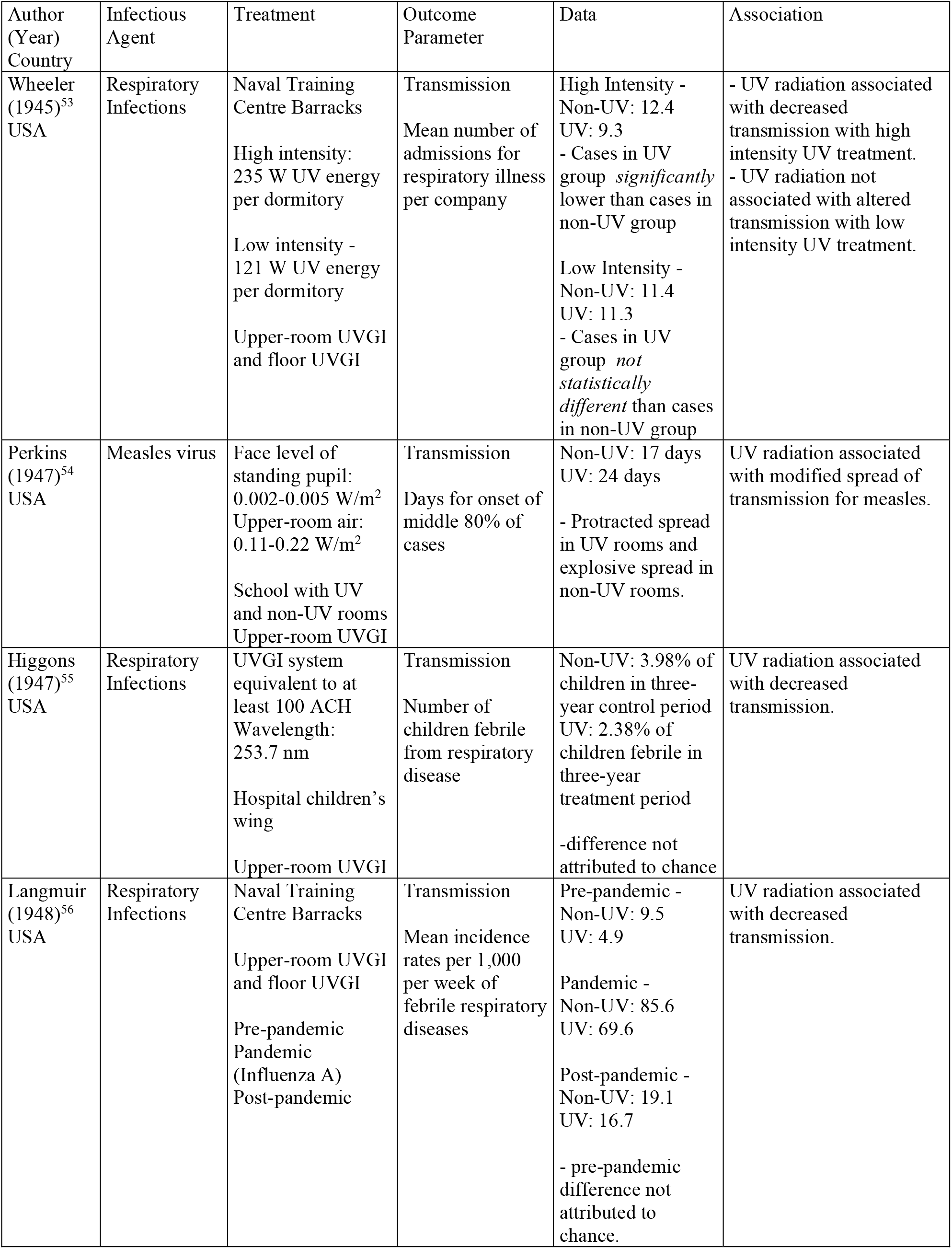
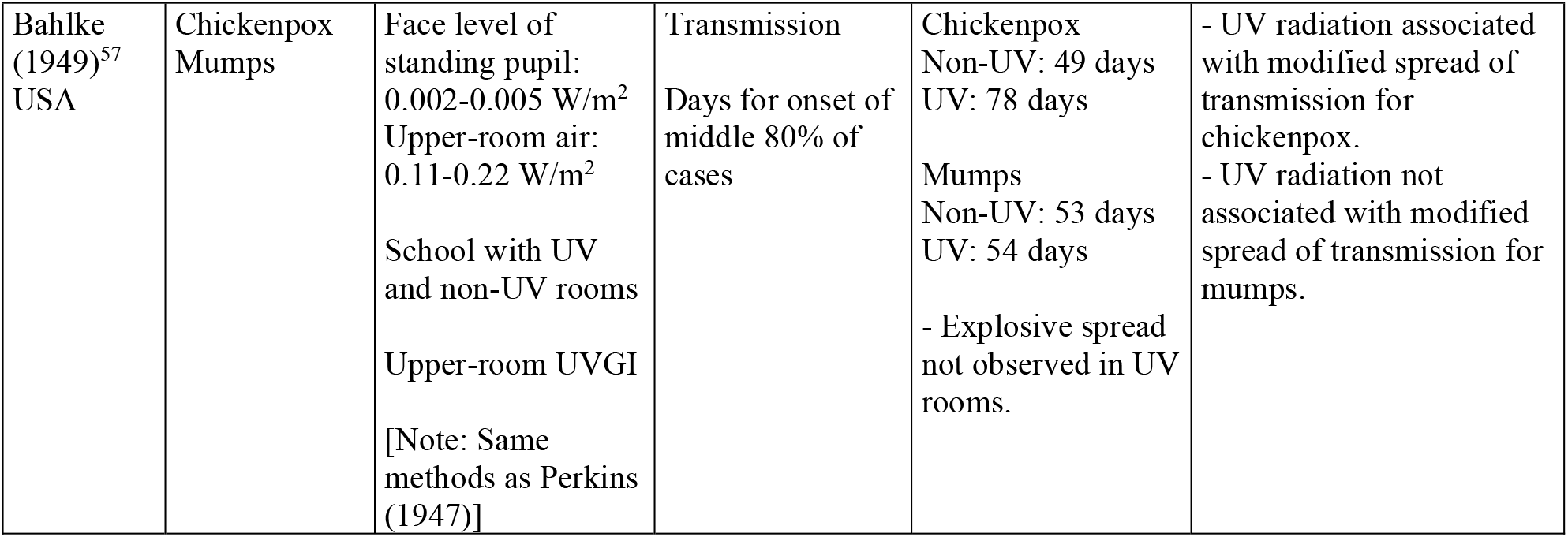
Summary of characteristics and findings for human studies of UV treatments.

| Author<br>(Year)<br>Country | Infectious<br>Agent | Treatment | Outcome<br>Parameter | Data | Association |
| --- | --- | --- | --- | --- | --- |
| Wheeler<br>(1945)<sup>53</sup><br>USA | Respiratory<br>Infections | Naval Training<br>Centre Barracks<br>High intensity:<br>235 W UV energy<br>per dormitory<br>Low intensity -<br>121 W UV energy<br>per dormitory<br>Upper-room UVGI<br>and floor UVGI | Transmission<br>Mean number of<br>admissions for<br>respiratory illness<br>per company | High Intensity -<br>Non-UV: 12.4<br>UV: 9.3<br>- Cases in UV<br>group *significantly*<br>lower than cases in<br>non-UV group<br>Low Intensity -<br>Non-UV: 11.4<br>UV: 11.3<br>- Cases in UV<br>group *not*<br>*statistically*<br>*different* than cases<br>in non-UV group | - UV radiation associated<br>with decreased<br>transmission with high<br>intensity UV treatment.<br>- UV radiation not<br>associated with altered<br>transmission with low<br>intensity UV treatment. |
| Perkins<br>(1947)<sup>54</sup><br>USA | Measles virus | Face level of<br>standing pupil:<br>0.002-0.005 W/m<sup>2</sup><br>Upper-room air:<br>0.11-0.22 W/m<sup>2</sup><br>School with UV<br>and non-UV rooms<br>Upper-room UVGI | Transmission<br>Days for onset of<br>middle 80% of<br>cases | Non-UV: 17 days<br>UV: 24 days<br>- Protracted spread<br>in UV rooms and<br>explosive spread in<br>non-UV rooms. | UV radiation associated<br>with modified spread of<br>transmission for measles. |
| Higgon's<br>(1947)<sup>55</sup><br>USA | Respiratory<br>Infections | UVGI system<br>equivalent to at<br>least 100 ACH<br>Wavelength:<br>253.7 nm<br>Hospital children's<br>wing<br>Upper-room UVGI | Transmission<br>Number of<br>children febrile<br>from respiratory<br>disease | Non-UV: 3.98% of<br>children in three-year control period<br>UV: 2.38% of<br>children febrile in<br>three-year<br>treatment period<br>-difference not<br>attributed to chance | UV radiation associated<br>with decreased<br>transmission. |
| Langmuir<br>(1948)<sup>56</sup><br>USA | Respiratory<br>Infections | Naval Training<br>Centre Barracks<br>Upper-room UVGI<br>and floor UVGI<br>Pre-pandemic<br>Pandemic<br>(Influenza A)<br>Post-pandemic | Transmission<br>Mean incidence<br>rates per 1,000<br>per week of<br>febrile respiratory<br>diseases | Pre-pandemic -<br>Non-UV: 9.5<br>UV: 4.9<br>Pandemic -<br>Non-UV: 85.6<br>UV: 69.6<br>Post-pandemic -<br>Non-UV: 19.1<br>UV: 16.7<br>- pre-pandemic<br>difference not<br>attributed to<br>chance. | UV radiation associated<br>with decreased<br>transmission. |
| Bahlke (1949)<sup>57</sup><br>USA | Chickenpox<br>Mumps | Face level of standing pupil:<br>0.002-0.005 W/m<sup>2</sup><br>Upper-room air:<br>0.11-0.22 W/m<sup>2</sup><br>School with UV and non-UV rooms<br>Upper-room UVGI<br>[Note: Same methods as Perkins (1947)] | Transmission<br>Days for onset of middle 80% of cases | Chickenpox<br>Non-UV: 49 days<br>UV: 78 days<br>Mumps<br>Non-UV: 53 days<br>UV: 54 days<br>- Explosive spread not observed in UV rooms. | - UV radiation associated with modified spread of transmission for chickenpox.<br>- UV radiation not associated with modified spread of transmission for mumps. |

### Aerosolized virus studies

Table 3 shows that 17 viruses and five bacteriophages from 16 studies were inactivated by UV radiation. Generally, susceptibility was determined from the dose-response relationship of UV dose and survival fraction^11,13,15,32-34^; however, Walker and Ko^10^ calculated susceptibility from a single dose and corresponding survival fraction, and Lin et al^35^ used both approaches. Some entries in Table 3 are presented as survival fraction calculated from the reported efficiency^36-38^ or the reported log reduction.^39^ For Qiao et al^39^ and Pearce-Walker et al,^39^ a lower detection limit was used to calculate the reported efficiency and log reduction, respectively. For Verreault et al,^40^ the reported relative infectious ratio appears to be equivalent to survival fraction; however, associations are presented with respect to relative infectious ratio. For 3 studies, UV radiation was associated with survival fraction.^37-39^

Increasing UV dose was associated with decreasing survival fraction for 10 viruses and five bacteriophages from 12 studies. Increasing UV dose was associated with decreasing survival fraction where the dose-response relationship was used to calculate susceptibility for seven studies (Table 3).^11,13,15,32-35^ Additionally, UV dose was associated with decreasing survival fraction where dose varied by exposure time,^36,40^ number of UV fixtures,^12^ and fluence rate.^41-42^

Increasing RH was associated with decreasing susceptibility in four studies^11,13,32-33^ for a variety of infectious agents including viruses (Influenza A, Vaccinia virus, PRRSV) and bacteriophages (MS2, phiX174, phi6, T7) (Figure 2). Cutler et al^33^ reported that PRRSV susceptibility was significantly lower at ≥80%RH compared with 25%RH-79%RH. In addition, four studies that report susceptibility at one RH are included in Figure 2^10,15,34-35^ where three viruses were coronaviruses (murine hepatitis virus (MHV) coronavirus, human coronavirus 229E, human coronavirus OC43). Considering the findings for influenza A^13,34^ and coronaviruses,^10,15^ UV radiation inactivated these enveloped, single-stranded RNA viruses and increasing UV dose was associated with decreasing survival fraction characterized by the susceptibility. If enveloped, single-stranded RNA animal viruses behave like influenza A (Figure 2), then increasing RH may be associated with decreasing susceptibility to UV radiation of coronavirus. The design of the UV radiation in an HVAC system should consider the reported coronavirus susceptibility recognizing that two are dose-response^15^ and one is single dose^10^ (Table 3).

**Figure 2.**
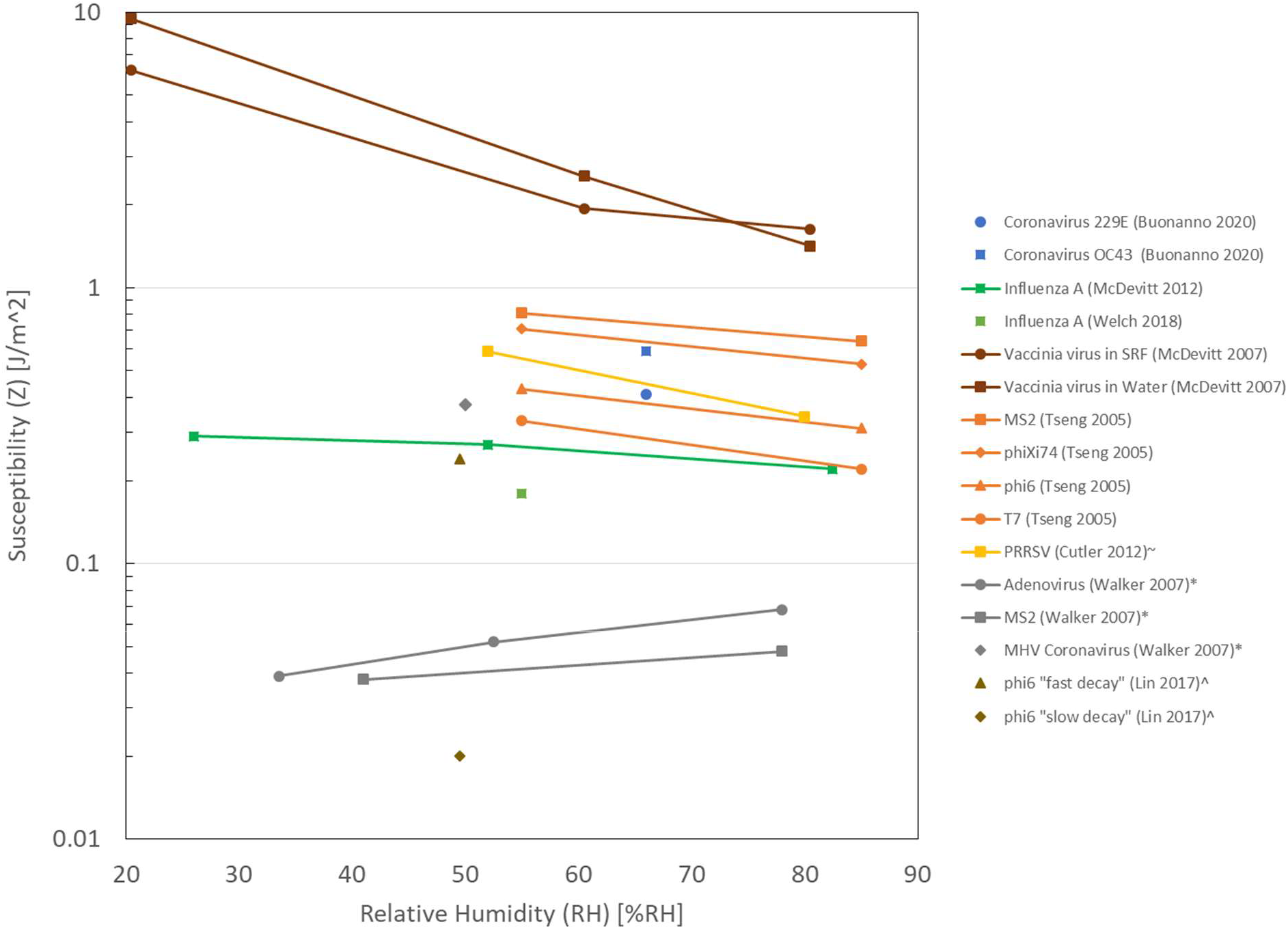
UV radiation susceptibility (Z) and relative humidity (RH) Each colour represents one study. Reported RH ranges shown as average RH values. ^∼^Cutler at al^33^ reported that susceptibility (Z) was significantly lower at ≥80%RH (shown at 80%RH) compared with 25%-79%RH (shown at 52%RH). ^*^Walker and Ko^10^ calculated susceptibility from a single dose and corresponding survival fraction, rather than dose-response of UV dose and survival fraction. ^^^Lin et al^35^ calculated susceptibility from a single dose and corresponding survival fraction for “fast decay” and from the dose-response of UV dose and survival fraction for “slow decay”.

Bacteriophage MS2 showed a discrepancy where Tseng and Li^32^ found that increasing RH was associated with decreasing susceptibility and Walker and Ko^10^ found that increasing RH was associated with increasing susceptibility. Walker and Ko^10^ acknowledged that this relationship for bacteriophages (MS2) and animal viruses (adenovirus) was different from that reported previously for bacteria. Other differences between the two studies of bacteriophage MS2 include the susceptibility calculation and suspending medium. Susceptibility was calculated using dose-response of UV dose and survival fraction by Tseng and Li^32^ and using a single dose and survival faction by Walker and Ko.^10^ Deionized water was used by Tseng and Li^32^ and phosphate buffered saline with 0.01% Tween 80 and Antifoam A was used by Walker and Ko.^10^

Three studies examined upper-room UVGI using a room-sized chamber.^12,41-42^ McDevitt et al (2008) examined the effect of summer conditions (80% RH and fan directing air downwards) and winter conditions (40% RH and fan directing air upwards) on survival of vaccinia virus, in addition to number of fixtures and ACH for upper-room UVGI. Comparing winter and summer conditions, increasing RH and changing fan direction were associated with increasing survival fraction and decreasing equivalent ACH. Overall, increasing RH was associated with increasing survival fraction, decreasing susceptibility and decreasing equivalent ACH. These findings suggest that the design of UV radiation in an upper-room UVGI system should consider the typical variation of indoor relative humidity throughout the year.

### Modelling studies

In the experimental study by McDevitt et al^12^ increasing ACH was associated with increasing survival fraction. Two modelling studies confirmed the association of increasing ACH and increasing survival fraction (Table 4).^43-44^ Increasing ACH is associated with increasing survival fraction because the increased ACH decreases the time that the infectious agent is exposed to UV radiation. Li et al^43^ considered the removal survival fraction which was the sum of the survival fraction attributed to UV radiation and the survival fraction attributed to ventilation. Increasing ACH was associated with decreasing removal survival fraction and decreasing ventilation survival fraction despite an increasing UV radiation survival fraction. The relationship between ACH and UV radiation is an important design consideration.

Susceptibility (Z), like those calculated in the aerosolized virus studies, are important input parameters in modelling studies.^43-45^ Three modelling studies used computational fluid dynamics (CFD) models (Table 4) to investigate the association of UV device location and UV dose and/or survival fraction. Li et al^43^ found that the survival fraction was decreased when the UV devices were located at the ceiling in the centre of the four walls compared with at the four corners. Sung and Kato^45^ found that the highest UV dose and lowest survival fraction were associated with the UV device being located opposite the one exhaust. Noakes et al^47^ found that UV dose was less affected by which one of the four UV devices was active when ventilated air was supplied at the floor and extracted at the ceiling compared with when ventilated air was supplied at the ceiling and extracted at the floor.

Furthermore, UV dose was associated with airflow pattern. Noakes et al^47^ found a higher average UV dose in the occupied region of the room when ventilated air was supplied at the floor and extracted at the ceiling compared with when ventilated air was supplied at the ceiling and extracted at the floor. The modelling of upper-room UVGI confirmed that designs must consider airflow pattern,^47^ ACH,^43-44^ and UV device location.^43,45,47^

UV radiation was associated with decreased attack rate which is the number of new cases in population at risk divided by number of persons at risk in population. Zheng et al^48^ found that attack rate decreased 87.8% when UVGI was modelled on a cruise ship where the ACH due to UV was 12 ACH.

### Animal and human studies

As early as 1936, UV radiation was associated with decreased influenza transmission in an animal model^49^ (Wells, 1936) (Table 5). UV radiation was associated with decreased virus transmission and infection incidence in three of the animal studies.^49-51^ Dee et al^52^ acknowledged that the lack of effect was likely due to insufficient exposure time to the UV radiation (Table 5).

All of the five human studies^53-57^ were from the 1940s and investigated upper-room UVGI (Table 6). UV radiation was associated with decreased transmission of respiratory infections in three studies.^53,55-56^ In the studies of barracks, Wheeler et al^53^ found that high intensity UV treatment was required to decrease transmission. UV radiation was associated with modified spread of transmission, but not prevention of transmission, of measles^54^ and chickenpox^57^ but not mumps.^57^

### Risk of bias

The risk of bias evaluation for the experimental studies demonstrated three scenarios (Table 7). Seventeen studies had low risk of bias for all three domains: selection bias, information bias, confounding. Seven studies had low risk of bias for selection bias and confounding but unclear risk of bias for information bias due to lack of clarity in the description of the UV radiation. Of these seven studies, four were aerosolized virus studies, two were animal studies, and one was a human study. One aerosolized virus study had low risk of bias for selection bias but high risk of bias for information bias and confounding due to lack of calibration of fluorescent bioaerosol count (FBC) and potential for UV radiation to affect fluorescence. Su et al^58^ cite other studies where FBC and cultures provided a predictable functional relationship which could be seen as a calibration of this potentially powerful and useful measurement tool; however, they do not provide such a relationship. Their FBC and culture data are not compared in a way that readers can clearly see how an FBC measure predicts a concentration of a pathogen in question. Su et al^58(p8)^ recognize that “[t]here is no available research about how UV light affects bioaerosols that generate a fluorescence signal.” The risk of bias evaluation for the seven modelling studies resulted in low risk of bias for all three domains (Table 8): definition, assumption, validation.

**Table 7.** Risk of Bias for Experimental Studies.

| Study | Selection Bias | Information Bias | Confounding* |
| --- | --- | --- | --- |
| *Aersolized Virus* |  |  |  |
| Jensen (1964)<sup>36</sup> | low | unclear | low |
| Tseng (2005)<sup>32</sup> | low | low | low |
| Walker (2007)<sup>10</sup> | low | low | low |
| McDevitt (2007)<sup>11</sup> | low | low | low |
| Su (2007)<sup>58</sup> | low | high | high |
| First (2007)<sup>41</sup> | low | low | low |
| McDevitt (2008)<sup>12</sup> | low | low | low |
| Terrier (2009)<sup>37</sup> | low | unclear | low |
| McDevitt (2012)<sup>13</sup> | low | low | low |
| Cutler (2012)<sup>33</sup> | low | low | low |
| Verreault (2015)<sup>40</sup> | low | unclear | low |
| Lin (2017)<sup>35</sup> | low | low | low |
| Welch (2018)<sup>34</sup> | low | low | low |
| Buonanno (2020)<sup>15</sup> | low | low | low |
| Pearce-Walker (2020)<sup>39</sup> | low | unclear | low |
| Qiao (2020)<sup>38</sup> | low | low | low |
| *Animal Studies* |  |  |  |
| Wells (1936)<sup>49</sup> | low | unclear | low |
| Jakab (1982)<sup>50</sup> | low | low | low |
| Dee (2006)<sup>52</sup> | low | unclear | low |
| Jaynes (2020)<sup>51</sup> | low | low | low |
| *Human Studies* |  |  |  |
| Wheeler (1945)<sup>53</sup> | low | low | low |
| Perkins (1947)<sup>54</sup> | low | low | low |
| Higgon (1947)<sup>55</sup> | low | low | low |
| Langmuir (1948)<sup>56</sup> | low | unclear | low |
| Bahlke (1949)<sup>57</sup> | low | low | low |

\* Confounding assessed for our comparison of interest.

**Table 8.** Risk of Bias for Modelling Studies.

| Study | Definition | Assumption | Validation |
| --- | --- | --- | --- |
| Noakes (2006)<sup>47</sup> | low | low | low |
| Rudnick (2007)<sup>42</sup> | low | low | low |
| Li (2010)<sup>43</sup> | low | low | low |
| Sung (2011)<sup>45</sup> | low | low | low |
| Zheng (2016)<sup>48</sup> | low | low | low |
| Firrantello (2018)<sup>46</sup> | low | low | low |
| Buchan (2020)<sup>44</sup> | low | low | low |

## Discussion

UV inactivation of airborne viruses is governed by the UV dose. The required dose varies depending on the type of virus, capsid structures, and host cell repair mechanisms^59^. Tseng and Li^32^ found that dsRNA and dsDNA viruses required a dose that was 2 times higher than their single strand counterparts for 90% inactivation. Walker and Ko^10^ came to a similar conclusion when they found that adenovirus was more resistant to inactivation compared to SARS. This resistance is attributed to the double stranded nature of its DNA genome and its ability to shield or consume UV radiation using small proteins concentrated along with viral particles^59^.

External factors such as ventilation and relative humidity also play an important role in UV effectiveness. Relative humidity is uniquely intertwined with UV inactivation. Many studies in this systematic review indicate that relative humidity has a marked, sometimes statistically significant effect on UV inactivation^10-13,33^. Further research must be done to ascertain the interaction of relative humidity and UV inactivation in a real-world context with a diverse list of infectious agents. The effect of ventilation on UV effectiveness is similarly complex. Increasing ventilation rate and UVGI are inversely related. At a higher ventilation rate infectious agents are removed from the space at a faster pace. This results in shorter exposure times thereby decreasing the effectiveness of the UVGI system^41^. Dee et al^52^ stated that in their experiments UVC provided no reduction in aerosol transmission of PRRSV due to insufficient exposure time. McDevitt et al^12^ argued that the combination of increased ventilation and upper room UVC is “more than merely additive”. The overall effect of a higher ventilation rate results in an increase in the ACH_uv_ (effective ventilation due to UVC). Li et al^43^ came to a similar conclusion that while UV disinfection efficiency decreases when ventilation rate increases, the overall infectious agent removal rate increases. In addition to ventilation rates, airflow patterns can have a meaningful impact on UVGI effectiveness^47^. Well mixed air allows UVGI to be more effective^12,41,47^. UVGI systems should be designed specifically for the targeted space. Ventilation rate, airflow patterns, and relative humidity should all be taken into consideration. In spaces with no ventilation systems or where increasing the ventilation rate would not be feasible, UVGI can provide cost effective air disinfection^60^. Spaces with a low outdoor air fraction can also benefit by adopting UVGI^50^. When considering upper room UVGI, lamp placement can have a significant impact on the effectiveness of the inactivation^47^. UV devices set on a wall rather than the corners of the room improves the effectiveness of the UVGI system^43^. Building owners and operators should consult an expert when examining the feasibility of installing UVGI in their space. First et al^41(p328)^ states that “upper-room UVGI must be approached as a carefully interdependent system with critical interactions among luminaire selection, luminaire placement, and all aspects of a ventilation system.”

When investigating the effectiveness of UVGI it is important to carefully consider the viral challenge and ensure the experiment mimics natural conditions. Even though Jensen^36(p420)^ used viral aerosol concentrations that were “many times greater than one would normally expect to encounter under natural conditions”, they stated that UVGI used in conjunction with filtration “should kill virtually all viruses”. Walker and Ko^10^ discussed the importance of aerosolization, sampling, and medium. Some viruses might be inactivated in the act of aerosolization or sampling; a medium with a high protein concentration might protect the targeted virus from inactivation^10^. UV susceptibility in liquid suspensions cannot be substituted for susceptibility in aerosols. Walker and Ko^10^ found that for the viruses they tested, UV susceptibility was higher in aerosols than in liquid suspensions. Attenuated strains of infectious agents can be used as a safer alternative to the actual virus as they are expected to closely mimic the behaviour of the actual viruses compared to an alternative surrogate^39^. With regards to safety, Welch et al^34^ and Buonanno et al^15^ have proposed the use of far UVC (222 nm) as a safer alternative to conventional UVGI (254 nm). It was demonstrated that far UVC has a similar inactivation efficiency to conventional UVGI for aerosolized coronavirus while not appearing to be cytotoxic to human cells and tissues in vitro or in vivo^15,61^.

Airborne transmission of respiratory pathogens is a serious issue. Like improving filtration and increasing ventilation, UVGI is a passive mitigation measure that can have an important impact on virus transmission. No responsibility is placed on the individuals in the space. Langmuir et al^56^ suggests that upper room UVGI by itself is not an adequate method of air disinfection. A well designed UVGI system should work in conjunction with other mitigation measures such as adequate filtration and ventilation^36,43,44,47,51^. Perkins^54^ and Bahlke^57^ found that even in the event of an outbreak, the use of UVGI led to low grade protracted epidemics of measles and chickenpox respectively, as opposed to large explosive episodes. The effectiveness of UVGI has been known for a long time. The benefits of a well designed UVGI system outweigh the principal and maintenance costs^55^. The time has come for UVGI to be considered as essential as ventilation and filtration.

The results of this systematic review revealed several important findings. First, viruses and bacteriophages were inactivated by UV radiation. Second, increasing UV dose was associated with decreasing survival fraction of viruses and bacteriophages. Third, increasing relative humidity was associated with decreasing susceptibility to UV radiation. Fourth, UV dose and corresponding survival fraction were affected by airflow pattern, ACH, and UV device location. Finally, UV radiation was associated with decreased transmission in both animal and human studies. While some of these findings may be well-established in the UV / technical literature, the value of this review is in bringing together this information in a comprehensive and rigorous manner to inform practical applications and set-up of UV systems in the built environment to assist with infection control. Further, we have identified gaps in the scientific literature that warrant attention to advance this important field.

Studies characterized as in-duct UVGI are designed with mechanically induced air flow with a controlled mean velocity which transports air through an irradiated zone inside a duct. In-duct UVGI lends itself to greater experimental control of the UV dose than the upper-room UVGI, although laminar duct flow, non-uniform velocity profiles, and radiation distribution always lead to some dose variance for in-duct systems. Also, in-duct configurations tend to simulate practical conditions where UVGI is installed in HVAC systems or air purifiers. Non-enclosed systems like upper-room UVGI depend on air circulation to drive the particles to an irradiated zone near the ceiling and does not require a controlled mean velocity: air moves through the UV zone generally due to air currents which are subject to room-scale turbulence. Thus their evaluation becomes more complex, needing to take into account ventilation configurations, air currents and lamp installation locations.

Two main thrusts of the research emerged: (1) research that focused on the effect of UV radiation on aerosolized virus survival and (2) research that considered some or all of the transmission chain from infected host to aerosolized virus to infected target where specific UV radiation configurations or scenarios were evaluated.

### Implications for research

Future research must ensure that UV dose and UV design requirements are clearly described within the context of their own study in order to simplify comparison between studies. A common metric used for quantifying the effects of UV on airborne pathogens was the measure of the survival fraction, comparing a quantity before and after the UV intervention. This metric is easy to understand but is highly dependent on the configuration of the system for exposing the aerosol to UV radiation. It shows how a system in its entirety kills or inactivates the pathogen in question, but does not isolate the more fundamental dose-response as the susceptibility (*Z*) measure, which is the exponent or linear slope on a semi-log Cartesian plot of the UV dose versus survival fraction. An encouraging and important trend evident in this body of work is the general evolution toward more rigorous control of experimental conditions which lend themselves to clear quantification of dose-response. Knowing that not all UV sources are alike, and that flux divergence, reflection, air velocity profile and lack of turbulence can lead to non-uniform UV radiation of aerosols allows researchers to focus on comparing susceptibility (*Z*) values, a single parameter. In some cases, the UV radiation and aerosolized virus studies were able to provide the mechanistic quantitative measure of susceptibility (*Z*) which is useful for cross study comparisons and is an important input parameter in modelling studies.

In addition to research examining the effect of UV radiation on aerosolized virus survival, the other main focus was research considering some or all of the transmission chain from infected host to aerosolized virus to infected target under specific UV radiation conditions. These results tend to be somewhat anecdotal because there is no standard test for the full transmission chain between two or more people sharing breathing space in the built environment. This is a common challenge in many fields of research. There would be value if the community moved to a more standardized test case configuration such as a standard room and ventilation system configuration, so that discrepancies attributable to factors such as geometry and flow field could be eliminated. The ASHRAE 185.1 standard states “Test standards form the foundation for air-cleaner selection in the ventilation industry. U.S. Environmental Protection Agency (USEPA) literature states that the most important need in the area of ultraviolet germicidal irradiation (UVGI) is industry standards to rate installed devices.”^62(p2)^

All human studies of upper-room UVGI that we identified were field studies conducted between 1945-1949. In a historical review, Reed^63^ indicates that UVGI fell out of favour after the 1940s due to inconsistent UV effects on measles transmission in schools, which were later attributed to measles exposure outside of schools. Additionally, no human studies on virus transmission and UV radiation were found after the 1940s which is a time period before vaccinations for measles,^64^ chickenpox,^65^ and mumps.^66^ Reed^63^ attributes more recent attention on UVGI to bacteria, tuberculosis, and viruses, influenza and SARS. In the current review, recent studies of UVGI are modelling studies in hospital settings from 2006 to 2020 in which influenza (2011) and coronavirus (2020) were considered. More field studies of upper-room UVGI are warranted to advance our understanding of its applicability.

### Implications for practice

The design of the UVGI for implementation in a building environment, whether in-duct or upper-room, should consider which virus is targeted. An additional consideration is how the susceptibility of that virus is affected by changes in relative humidity (Figure 2). If more than one infectious agent is targeted, then a range of susceptibilities should be considered. Jaynes et al^51^ designed their UVGI system to target one bacteria and two viruses (Table 5). Practical application of UVGI systems should take into account lessons learned from modelling studies; i.e. UV dose produced by the UV system may be affected by the airflow pattern, ACH and UV device location.^43-45,47^

Generally, in-duct UVGI can be used to prevent in-building transmission when ventilation supply air has some fraction which is recycled, a practice which is a common means to reduce heating/cooling costs. Also, in-duct UVGI could be incorporated into a ducted air purification system which exhausts back into the source space directly and might be a tool used to remove airborne virus in a space at a rate higher than could be achieved by the building air handling system alone. In both of these cases the survival of pathogen after UV exposure would need to be extremely low for them to be effective (though in practice in-duct systems would generally operate in series with aerosol removal by filtration). A consensus on an acceptable standard for virus reduction due to UV treatment might be helpful. Experimental studies may benefit from using the ASHRAE Standard 52.2 test duct which has been used in recent filtration studies.^67-68^ Simply seeing more studies recognize that UV radiation dose is dependent on the radiant flux, which is often not uniform, shows a positive trend. For ducted systems, flow that is well-mixed and close to uniform in velocity profile is helpful in evenly dosing aerosols with UV.

Upper-room UVGI is specifically designed to reduce the buildup of pathogen in the shared breathing space of occupants and in this technology there is definitely no standard for what is an acceptable performance for these systems. As was done by First et al,^41^ Rudnick and First,^42^ and McDevitt et al,^12^ the equivalent ACH does seem to be a sensible way to calibrate these systems.

Two experimental studies^15,34^ and one modelling study^44^ investigated the effects of far-UVC light (222 nm) on virus survival as an alternative to conventional UVC light sources. Viruses used included human coronaviruses alpha HCoV-229E and beta HVCo-OC43,^15^ SARS-CoV-2,^44^ and Influenza A (H1N1).^34^ While conventional UV has been shown to reduce virus survival fractions, conventional UV can be carcinogenic and cataractogenic^34,44^ and a health hazard when exposed directly.^15^ Far-UVC light has been posited as an option for UV radiation as, since far-UVC light has a lower range “of less than a few micrometers, and thus it cannot reach living human cells in the skin or eyes,” the range is still greater than that of viruses, allowing UVC light to “penetrate and kill them.”^15(p1)^ Given concerns of ozone generating by 185nm UV, Welch et al^34^ measured O_3_ concentration and could not detect ozone with their 5ppb threshold of detection.

## Conclusion

This review provides a comprehensive and rigorous synthesis of the existing scientific literature examining the effectiveness of UV radiation and virus survival and transmission. Experimental studies of UV radiation have consistently demonstrated high susceptibility of viruses (or simulant agents) with sufficient UV dose. At this time, the UV susceptibility of aerosolized SARS-CoV-2 has yet to be reported. However, there are few studies examining the effect of UV radiation outside laboratory or simulated settings. Further, future field studies of real-world implementations of UVGI need to take into account the various factors that exist within ventilated indoor spaces that may modify UV effectiveness, including humidity, airflow pattern, air changes per hour, and UV device location. Research is needed to provide evidence of the effect of UV radiation along the chain of transmission in non-simulated “real life” settings.

## Data Availability

all data come from publicly available publications

## Acknowledgements

We thank Tara Landry and Alison Henry for conducting the peer review of the search strategies. We thank Samuel Ducholke, Kristen Rumbold, Larry Zhong, and Stella Mathews for their involvement in screening studies for inclusion.

